# Diagnostic Utility of Exome Sequencing for Polycystic Kidney Disease

**DOI:** 10.1101/2022.02.01.22269973

**Authors:** Alexander R. Chang, Bryn Moore, Jonathan Z. Luo, Gino Sartori, Brian Fang, Steven Jacobs, Yoosif Abdalla, Mohammed Taher, Regeneron Genetics Center, David J. Carey, William Triffo, Gurmukteshwar Singh, Tooraj Mirshahi

## Abstract

**Importance:** Most studies of ADPKD genetics have used select cohorts, focusing on *PKD1* and *PKD2* and more recently several other cystic genes. However, the population prevalence of ADPKD and the contribution of each cystic gene to ADPKD are not well understood.

**Objective:** Determine the prevalence of ADPKD, contribution of *PKD1, PKD2*, and other cystic genes to ADPKD in a large, unselected cohort.

**Design, Setting, and Participants:** We determined the prevalence of ADPKD In an unselected health system-based cohort of 173,954 subjects with the existing exome sequencing and extensive electronic health records, including abdominal imaging. The presence of genetic variants in *PKD1, PKD2*, and eleven other cystic genes was evaluated. Rare genetic variants were identified in patients with chart review confirmed diagnosis of ADPKD.

**Main Outcomes:** Diagnosis of ADPKD and presences of rare (AF<0.0001) missense, protein-truncating variants (PTVs), or copy number variants deletions (CNV) in *PKD1, PKD2*, or PTVs and CNVs in the following 11 genes: *ALG8, ALG9, DNAJB11, GANAB, HNF1B, IFT140, LRP5, PKHD1, PRKCSH, SEC61B*, and *SEC63*.

**Results:** Among 173,954 patients, there were 235 patients with chart review confirmed ADPKD (0.135%). Among patients with PTV or CNV in *PKD1*, 66/70 (94.2%) had ADPKD and 43/44 (97.7%) of patients with PTV or CNV in *PKD2* had ADPKD. In contrast, only 24/77 (31.2%) patients with a PKD1 missense variant previously classified as “likely pathogenic” had ADPKD. A rare variant was identified in a cystic gene in 180/235 (76.6%) of ADPKD patients, with the most common genes implicated *PKD1* (127) and *PKD2* (34) and then *IFT140* (7), *PKHD1* (3), *GANAB* (4), *HNF1B* (2), *ALG8* (1), *ALG9* (1), *IFT140*+*PKHD1* (1). The yield for a genetic determinant of ADPKD was 91.3% among those with a family history compared to 50.6% among those without a family history (p<0.0001). We report several previously unreported variants where pedigree data suggest pathogenicity. Atypical cystic genes *ALG8, ALG9, GANAB, HNF1B, IFT140*, and *SEC63* were associated with having any kidney or liver cystic ICD code, but not diagnosed ADPKD.

**Conclusions:** Exome sequencing established the molecular diagnosis for the vast majority of patients with ADPKD, revealed a wider range of ADPKD with atypical cystic genes. Additional population-based research cohorts are needed to carefully curate missense *PKD1* variants and variants in atypical cystic genes.

## Introduction

Autosomal dominant polycystic kidney disease (ADPKD) is the most common genetic cause of kidney failure. Prevalence estimates range from 3.96 per 10,000 for clinically-determined ADPKD (1) to 1 per 1000 in large cohorts with whole genome sequencing(2). The most common genetic variants that cause ADPKD are in *PKD1* (77%) and *PKD2* (15%), which encode polycystin-1 and polycystin-2, respectively(3). Milder cases of ADPKD without family history are often observed in clinical practice and are not well-represented in research cohorts. As studies of other Mendelian diseases have demonstrated, the population penetrance and phenotypic severity of ADPKD may be lower than those described in disease-specific cohorts, which tend to be overrepresented by patients with more severe presentations (4-6). An analysis of the Toronto Genetic Epidemiology of Polycystic Kidney Disease study reported that roughly one half of participants without family history of ADPKD do not have a detectable *PKD1* or *PKD2* variant(7). Some of these unexplained cases may be due to variants in other genes (e.g., *ALG9, GANAB, DNAJB11, HNF1B) (8-10)*. However, the penetrance and expressivity of variants in these atypical PKD genes remains largely unexplored.

Exome sequencing allows for exploration of all coding variants in a population to better understand penetrance and expressivity for rare diseases. For some genes where prior knowledge of genetic variants can lead to preventive clinical action, a genotype-based approach can be used to inform participants of their genetic findings(11). Despite recognition of high penetrance of pathogenic variants in *PKD1 and PKD2*, especially truncating *PKD1* variants(12, 13), a primarily genotype-based approach for early ADPKD has not yet been implemented partially due difficulty in capturing reliable *PKD1* variants from exome sequencing (14).

Early recognition of ADPKD may be important as an approved treatment exists(15), and there are several ongoing clinical trials testing new therapeutic options(16). Currently, diagnosis of ADPKD for most at-risk patients with family history of ADPKD is done by abdominal imaging(17). Genetic testing is generally only performed when ADPKD is suspected in persons without family history or with equivocal kidney imaging findings, in the setting of related kidney donation, or for family planning purposes(18). As NGS panels and exome sequencing are increasingly available for multiple medical conditions, there is an urgent need to better understand the significance of protein-truncating variants (PTVs) in atypical cystic disease genes and clarify the clinical significance of missense variants in *PKD1* and *PKD2* to improve counseling and guide management decisions.

In this study, we examine the utility of exome sequencing for deciphering monogenic causes of ADPKD in a large health-system based cohort including many patients without family history. We use a genotype-based approach to evaluate the prevalence of ADPKD and kidney and liver cyst diagnoses in PTV carriers of *PKD1, PKD2, ALG8, ALG9, DNAJB11, GANAB, HNF1B, IFT140, LRP5, PKHD1, PRKCSH, SEC61B*, and *SEC63*. In parallel, we identified clinically-confirmed ADPKD cases in a largely unselected, health system-based research cohort gaining insight into true population prevalence and the genetic causes of ADPKD in these patients.

## Methods

### Study Population

The Geisinger Institutional Review Board approved this study. Informed consent was waived as participants were previously consented in the MyCode(tm) Community Health Initiative and were studied as part of the Geisinger-Regeneron DiscovEHR collaboration(19). For this study we included 173,954 participants from the MyCode cohort who had exome sequencing and health record data available.

### Exome sequencing

Exome sequencing was performed in collaboration with Regeneron Genetics Center as previously described (19) (Supplemental Methods). Coverage depth was sufficient to provide more than 90% coverage of the targeted bases for 99% of samples. Alignments and variant calling were based on GRCh38 human genome reference sequence. Copy number variants for *PKD1, PKD2*, and atypical ADPKD genes were estimated using lattice-aligned mixture models (CLAMMS) software(20).

Nonsense or protein truncating variants (PTVs) in *PKD1, PKD2*, and 11 atypical cystic genes (*ALG8, ALG9, DNAJB11, GANAB, HNF1B, IFT140, LRP5, PKHD, PRKCSH, SEC61B, SEC63)* were included that had GQ score >20, AD_ALT>3, AD_ALT/AD_REF ≥0.5, and MAF<0.01. Inframe deletions and missense *PKD1* and *PKD2* variants with MAF <0.0001 listed in the Mayo PKD database (PKDB; http://pkdb.pkdcure.org), as Likely Pathogenic were included using the same criteria. We cross-referenced all variants with ClinVar interpretation and stars rating system (21), the PKDB (22), and Varsome, a database that considers case-control, computational, functional, and family segregation data to provide American College of Medical Genetics (ACMG)-guided classification of variants’ pathogenicity(21). For patients with clinical diagnosis of ADPKD, we also cataloged any accompanying rare (MAF <0.001) missense variants in *PKD1 and PKD2* and rare pLOF or missense variants in the 11 atypical cystic genes.

### Phenotyping using Electronic Health Record (EHR)

We used data from the EHR to identify participants with International Classification Diseases (ICD) diagnosis codes (Q61.2, Q61.3, 753.13, 753.12). Additional diagnoses examined included other cystic kidney diseases (Q61.5, Q61.8, Q61.9, 753.10), acquired kidney cyst (N28.1, 593.2), congenital kidney cyst (Q61.00, Q61.01, Q61.02, 753.11, 753.19), or liver cystic disease (Q44.6, 573.8). Based on our and others’ prior studies examining *ALG9* and *DNAJB11(8, 23)*, we anticipated that ADPKD and PLD diagnoses would be rare among carriers of atypical PTVs and therefore used a broader composite outcome of any kidney/liver cyst ICD code, which included ADPKD, cystic kidney disease, acquired renal cyst, and liver cyst diagnoses.

### Chart Reviews

Additional chart review was performed on select participant groups by at least 1 nephrologist with focus on kidney and liver imaging, nephrolithiasis, cerebral aneurysms, history of dialysis, transplant, family history of ADPKD or cerebral aneurysms, and clinical genetic testing. Blinded review of imaging was done by at least 1 radiologist with additional review by a second radiologist for questionable cases, and cases were discussed until consensus was achieved. Imaging phenotypes were classified as:

1. Typical; bilateral and diffuse distribution of cysts, with moderate or severe replacement of kidney tissue by cysts, where all cysts contribute similarly to total kidney volume, (for example see **Supplemental Figure 1**)
2. Mild; if there was mild bilateral/diffuse replacement of kidney tissue by cysts (for example see **Supplemental Figure 2**).
3. Atypical per Mayo clinic imaging classification (24) (for example see **Supplemental Figure 3**).

If patients lacked imaging that was available for radiologist review, we considered them to have typical ADPKD if they had consistent clinical history (e.g. ESKD due to ADPKD, imaging report and family history consistent with Ravine-Pei criteria) (17, 25, 26).

### Statistical Analyses

Firth logistic regression was used to assess the association of PTVs with ADPKD or any kidney or liver cysts determined by ICD code. Models were adjusted for age, sex, year of first outpatient encounter at Geisinger, and genetically-determined ancestry. First and second-degree relatives were removed in this analysis.

### Construction of patient pedigrees

After identifying first- and second-degree relatives of each participant with ADPKD using PRIMUS(27) and EHR-documented family history, we reconstructed pedigrees of patients with ADPKD. We then reviewed charts of family members of ADPKD patients to evaluate evidence for co-segregation of the variants in question with disease.

## Results

### Genotype-first analyses for pLOFs in *PKD1* and *PKD2*

From exome sequencing data of 173,954 MyCode participants, we identified 3,556 variants in *PKD1* (AF range 0.19 to -2.88×10^−6^), including 1,919 variants that are listed in gnomAD exome sequencing data and 1,842 listed in gnomAD genome sequencing data. ClinVar interpretation is available for 460 of the variants identified. Using ACMG criteria as assessed by VarSome, *<u>PKD1</u>* variants were classified as 58 benign, 667 likely benign (LB), 2,265 Variant of Uncertain Significance (VUS), 384 Likely Pathogenic (LP), and 137 as Pathogenic. For *PKD2*, we found 434 variants (AF range 0.36-2.88×10^−6^) with 177 variants present in single patients. Among the *PKD2* variants, 216 are in gnomAD exome sequence data and 204 are in gnomAD genome sequence data. Variants were classified using ACMG criteria per Varsome as 7 benign, 39 LB, 339 VUS, 19 LP, and 29 pathogenic. ClinVar had pathogenic or LP with two-star review status classification for only 16 *PKD1* and 7 *PKD2* variants found in our cohort.

We identified 135 individuals with PTVs in *PKD1* or *PKD2*, which include early terminations, frameshift due to indels, and splice site variants, as well as large chromosomal deletions (CNVs) that encompassed *PKD1* or *PKD2* (**Figure 1A**).

**Figure 1.**
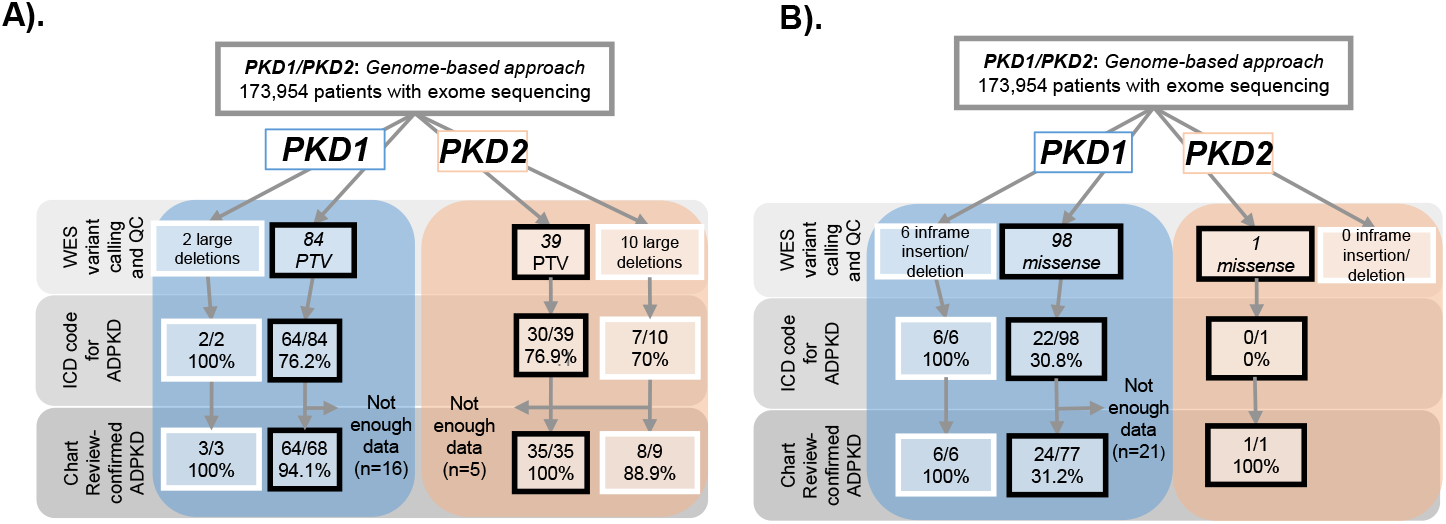
Genotype first approach-. Carriers of *PKD1* or *PKD2* A) CNV/PTVs or B) Inframe deletions/missense variants classified as likely pathogenic in PKDB. 173,954 participants with whole exome sequencing were evaluated for copy number variants (CNVs) or protein truncating variants (PTVs) in *PKD1* and *PKD2*. Electronic health records (EHR) of carriers of these variants were examined for ADPKD ICD codes (Q61.2, Q61.3, 751.12, 751.13), and chart reviewed to confirm presence of ADPKD.

### ADPKD diagnosis among *PKD1* PTV and CNV

Based on ICD code diagnoses, 2/2 (100%) patients with large genomic deletions in *PKD1* had ADPKD and 64/84 (76.1%) with *PKD1* PTVs had ADPKD. Among 68 patients with a *PKD1* PTV and sufficient data for chart and imaging review, 64 had ADPKD (94.1%). Four individuals with *PKD1* PTVs had imaging data sufficient to rule out ADPKD, and had no recorded family history of ADPKD. All participants with ADPKD and available imaging data had typical ADPKD imaging phenotype, with moderate-severe replacement of kidney tissue with cysts in a bilateral and diffuse distribution.

### ADPKD diagnosis among *PKD2* PTV and CNV

For *PKD2*, 7 out of 10 patients (70%) with large genomic deletions, and 30/39 (77%) patients with PTVs had ADPKD ICD codes **(Figure 1A)**. Chart review revealed that 4 individuals with *PKD2* PTVs and 1 with *PKD2* CNV, were under the age of 40 and lacked sufficient clinical or imaging information to make a determination about ADPKD. After excluding these individuals lacking data, 8 of 9 (88.9%) patients with large genomic deletions in *PKD2*, had ADPKD, and 35 of 35 (100%) patients with *PKD2* PTVs had ADPKD. *PKD2* variant carriers showed a wider spectrum of ADPKD severity, with 74% showing typical ADPKD, 17% with mild PKD, and 9% with atypical PKD.

### ADPKD diagnosis among carriers of known *PKD1* and *PKD2 in-frame deletions or* missense variants

**Figure 1B** shows the prevalence of ADPKD among 105 carriers of known rare in-frame deletions or missense variants in *PKD1* or *PKD2* listed in the PKDB. Chart review revealed that 21 patients had insufficient data to diagnose ADPKD. After removing these 21 patients, 24/77 (31.2%) with a *PKD1* LP variant, 6/6 with in-frame deletions. The one patient with a *PKD2* LP variant had ADPKD per chart review but did not have an ADPKD ICD code.

### Genotype-first analyses for *ALG8, ALG9, DNAJB11, GANAB, HNF1B, IFT40, LRP5, PKHD, PRKCSH, SEC61B, SEC63*

There were 3,793 patients with rare heterozygous PTVs and 61 CNV carriers in these 11 genes. Only 1/14 *GANAB*, 2/522 *PKHD1, 1/16 HNF1B, and 5/205 IFT40* PTV carriers had ADPKD ICD diagnosis. **Table 1** shows data comparing the prevalence of ADPKD ICD diagnosis and any kidney/liver cyst ICD diagnosis among PTV and CNV deletion carriers for these genes as well as *PKD1* and *PKD2*. As expected, having a PTV or CNV deletion in *PKD1* or *PKD2* was strongly associated with ADPKD diagnosis. For PTV or CNV deletion in the atypical cystic genes, *GANAB, IFT40, HNF1B* and *PKHD1* were significantly associated with ADPKD diagnosis. PTV or CNV deletion variants in *ALG8, ALG9, GANAB, HNF1B*, and *SEC63* were significantly associated with any kidney or liver cyst ICD diagnosis. Only two *17q12* deletion carriers (that encompasses *HNF1B*) and two *PKHD1* CNV deletion carriers had ADPKD diagnosis.

**Table 1.**
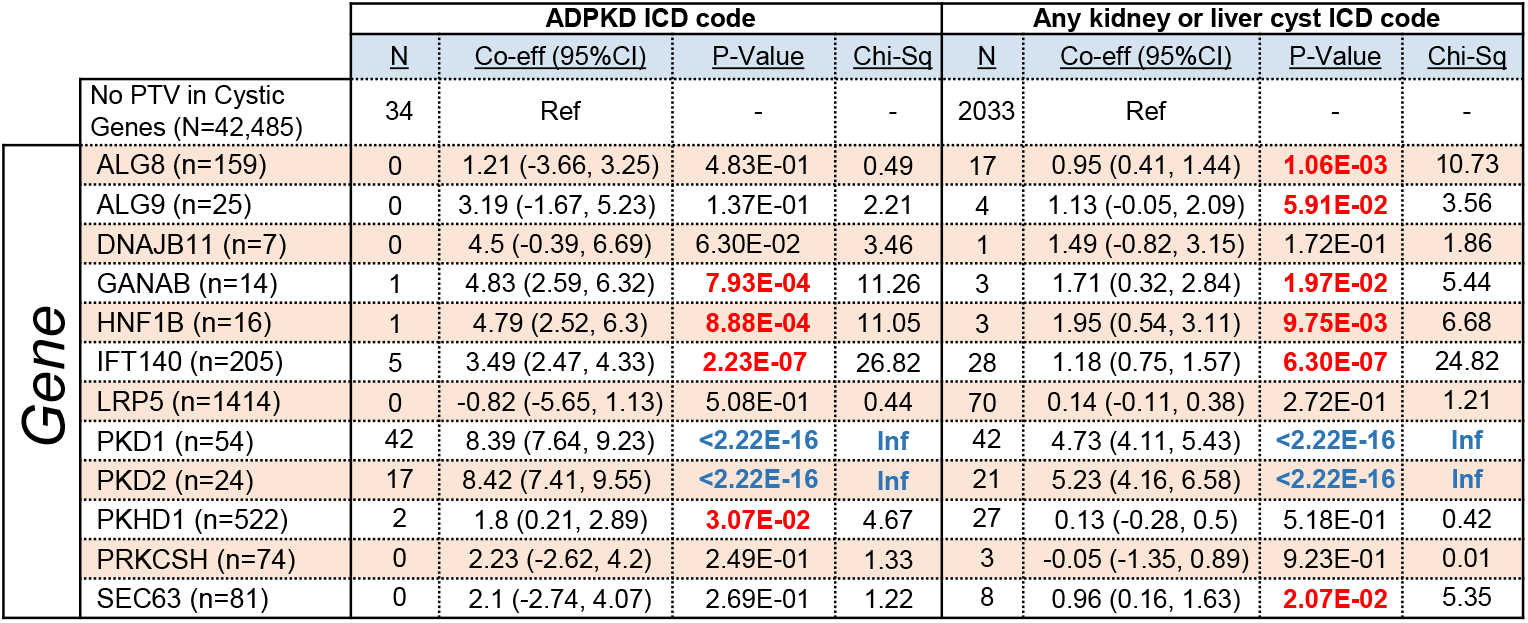
Risk of ADPKD or kidney/liver cyst ICD codes among those with PTV or CNVs in atypical APDPKD genes *ALG8, ALG9, GANAB, HNF1B, IFT40*, and *SEC63* are increased. Firth’s logistic regression was used to assess the association of PTV and CNVs with ADPKD and kidney/liver cyst determined by ICD code. The regression was adjusted for age, sex, year of first outpatient encounter at clinic, and genetically-determined ancestry. For these analyses, first- or second-degree relatives in the cohort were removed. For each gene, the logistic regression coefficient represents the expected change in log-odds of the outcome occurring if a PTV is present in the gene. A larger coefficient may be interpreted as a larger increase in the relative risk of the outcome (see statistical methods for regression details).

### Prevalence and phenotypic landscape of ADPKD

A total of 303 of 173,954 participants had at least 1 ADPKD ICD diagnosis. After excluding 42 who did not have ADPKD or were missing sufficient information for diagnosis, there were 235 participants with ADPKD and 26 who had phenotypes more closely matching other kidney diseases (**Figure 2A, B, Supplemental Table 1)**. Overall, a rare variant in one of the 13 cystic genes was identified in 180/235 (76.6%) of patients with ADPKD. When stratified by typical, mild, and atypical ADPKD, rare variants were identified in 81.1% for typical ADPKD, 68.8% for mild ADPKD, and 47.8% for atypical ADPKD. Stratified by EHR-reported family history, rare variants were identified in 91.3% with family history and 50.6% without family history.

**Figure 2.**
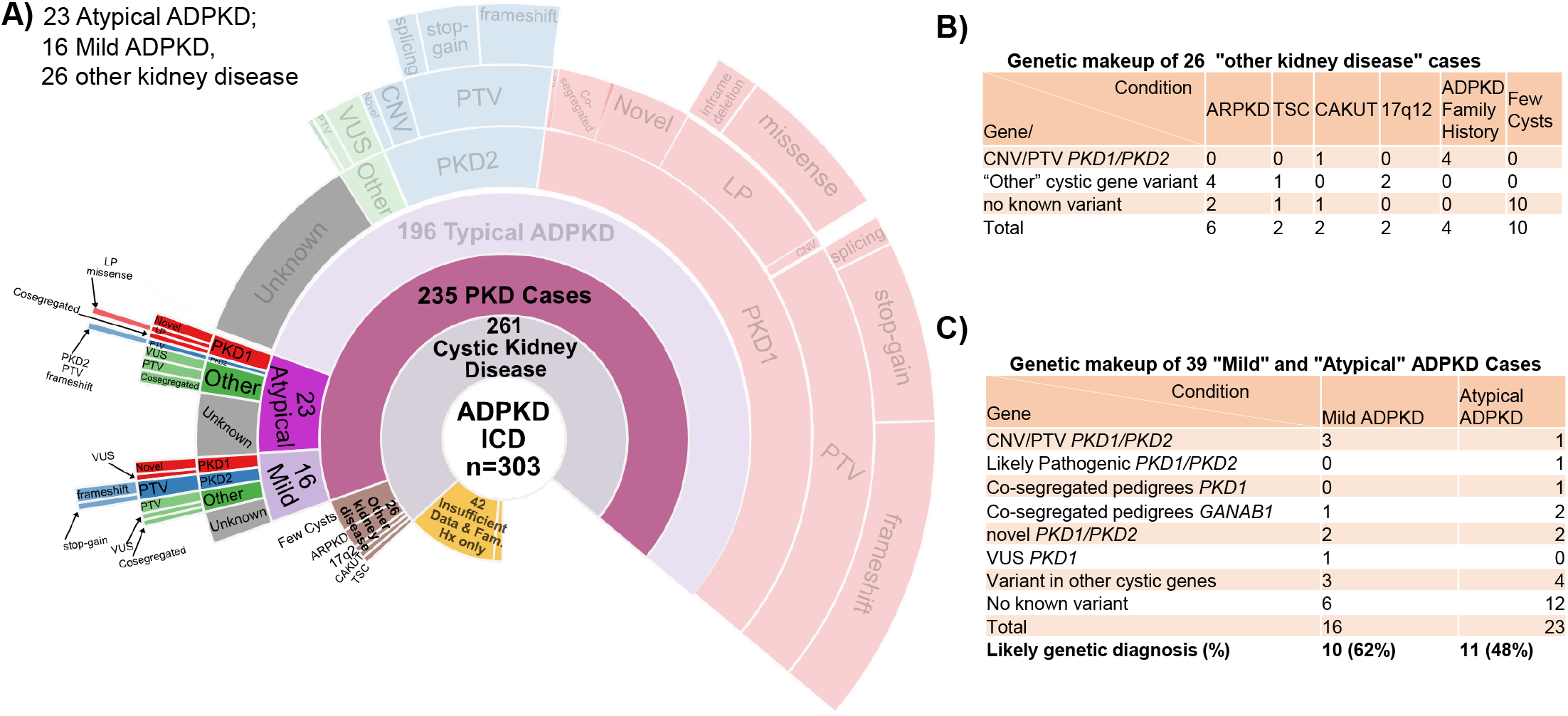
Phenotype first approach. Prevalence, phenotypic spectrum, and genetic determinants of ADPKD. **A)** Sunburst plot of various phenotypes in the EHR. A total of 173,954 subjects with whole exome sequencing were evaluated for ADPKD ICD codes (Q61.2, Q61.3, 751.12, 751.13), followed by chart review of 303 of them. Among these, 42 did not have enough data to diagnose any kidney disease. Of the remaining 261, 26 had other kidney diseases. From the remaining 235, Atypical and Mil ADPKD account for 23 and 16 patients respectively. The final 196 patients had typical ADPKD. Definition of each phenotype is detailed in the Methods. **B)** Of the 26 people who had other kidney diseases, 6 had ARPKD, 2 had TSC, 2 had CAKUT, 2 had 17q12 syndrome, and 4 had a family history of ADPKD and 10 only had “few cysts”. **C)** Individuals with atypical or mild ADPKD are much less likely to have a potential variant identified as a contributor to their ADPKD with only over half of each group being genetically resolved. Among these few had PKD1 or PKD2 PTVs or LPs, while the majority had novel variants in PKD1, PKD2 or in other cystic genes.

### Typical, Mild and Atypical ADPKD

We stratified the participants with ADPKD into 3 groups, typical, mild, and atypical as defined in the methods above. Among 235 ADPKD subjects we found: 196 typical (moderate/severe replacement of kidney tissue with bilateral/diffuse cysts), 16 mild (mild replacement of kidney tissue bilateral/diffuse cysts), and 23 atypical (**Figure 2A**,). Genetic variants in patients with mild or atypical ADPKD include those in *PKD1, PKD2* as well as other putative PKD genes such as *GANAB* **(Figure 2C)**. Of 39 participants with mild or atypical ADPKD, 18 (46%) did not have a candidate variant identified.

### *PKD1*/2 PTVs and CNVs account for most of ADPKD cases

Of the 196 patients with typical ADPKD, 99 had PTVs or CNVs in *PKD1* or *PKD2*. Specifically, 69 had a *PKD1* PTV [large deletion, stop gain, frame shift, splices site variant], 2 had chromosomal deletions encompassing *PKD1*, 23 had a *PKD2* PTV, and 5 had large chromosomal deletions encompassing *PKD2* (**Figure 3A**). Pedigree analyses were completed for most of the cases with typical ADPKD and a PTV/CNV for *PKD1* or *PKD2*. **Figure 3B** is an example of pedigree analysis for carriers of the protein truncating variant *PKD1_Glu1061Ter*, where the variant perfectly segregates with ADPKD; a kidney CT scan image from a variant carrier is also shown. Self-reported family history was much more common in individuals with typical ADPKD (60.2%), compared to atypical and mild ADPKD. Among the 196 typical ADPKD participants, 118 (60.2%) had self-reported family history of ADPKD with 110/118 (93.2%) having a variant in *PKD1* or *PKD2*.

**Figure 3.**
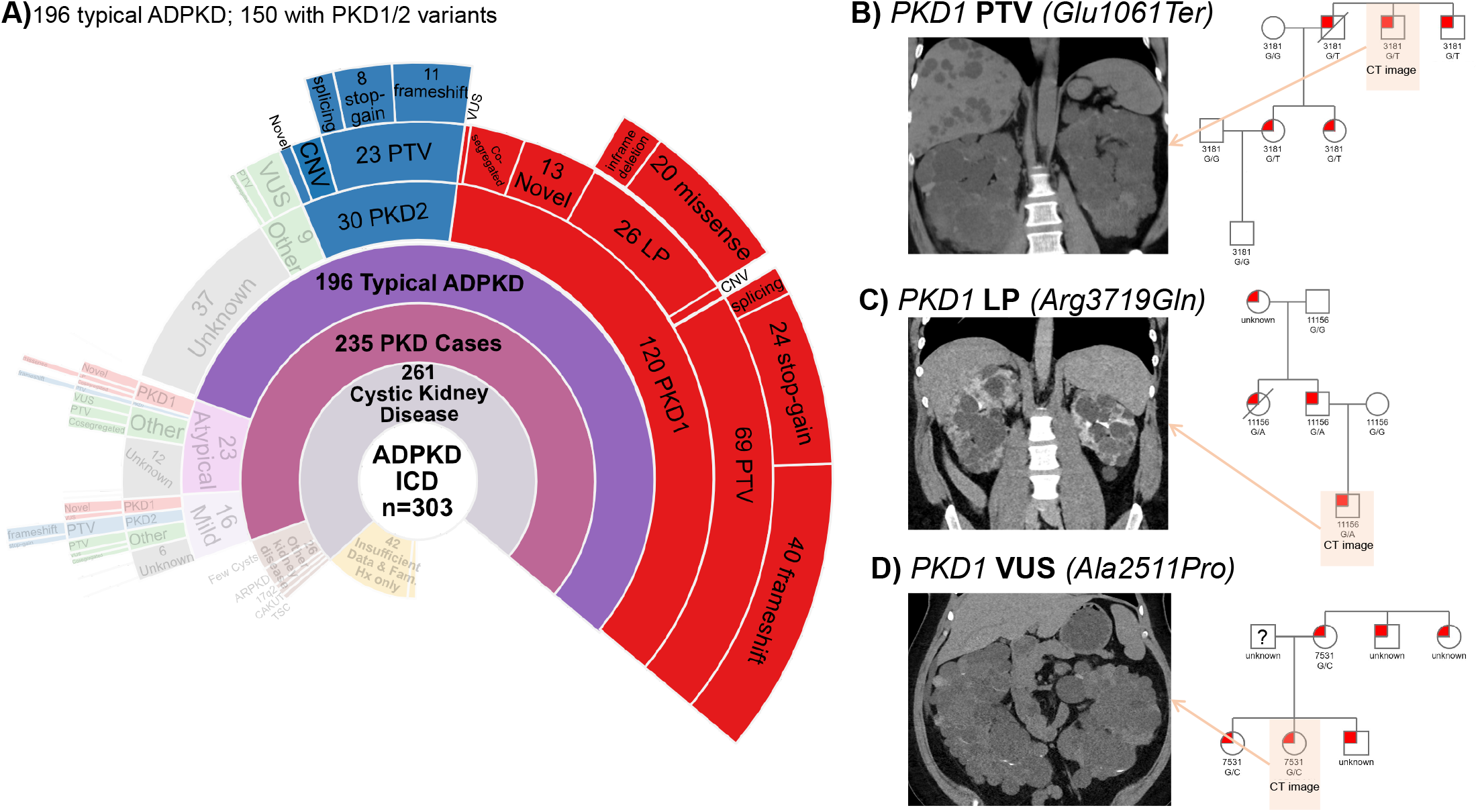
PKD1 and PKD2 variants among patients with typical ADPKD. **A)** Sunburst plot highlighting 150 typical ADPKD patients for whom a PKD1 or PKD2 variants were found. PKD1 variants comprise 80% of cases (120/150). **B)** Most variant identified were PTVs. Abdominal imaging for a patient with *PKD1(Glu1061Ter)* is shown, indicating large cystic kidneys, cysts in the liver. Family pedigree shows segregation of ADPKD among members with available genetic and clinical data. **C)** LPs in PKD1 were found in 26 patients but no PKD2 LPs were found in the cohort. Abdominal CT in the PKD1(Arg3719Gln) carrier shows presence of bilateral large cysts. Family pedigree shows ADPKD among variant carriers. **D)** A subset of typical ADPKD patient had variants n PKD1 or PKD2 that were previously unreported or had been classified as benign or VUS. Using family pedigrees, we confidently assigned genetic variants to 29 of these cases. CT image for carrier of *PKD1(Arg2511Pro)* which was been previously classified as likely benign by PKDB and a VUS by VarSome. The patient has large bilateral cystic kidneys, and the variant clearly segregates with ADPKD in pedigree analysis.

### Identifying known missense *PKD1*/*PKD2* variants in typical ADPKD

We identified 26 carriers of *PKD1* in-frame deletion or missense variants (MAF<0.0001) that are classified in PKDB as likely pathogenic (LP), and no carriers for *PKD2*. **Figure 3C** shows a pedigree analysis of a family with the LP *PKD1_Arg3719Gln* variant that segregates with ADPKD, along with a family member’s abdominal CT scan at age 25-30 showing typical ADPKD.

### Using family and cohort data to classify VUS and novel missense variants

We identified 25 ADPKD patients who had *PKD1* or *PKD2* variants that were reported as VUS or likely benign in PKDB or were not reported in that database (**Figure 3A**). The list of these patients and their variants in shown in **Supplemental Table 2**. For 9 of these patients there was sufficient clinical data from the EHR to confirm the presence of ADPKD, strongly suggesting these variants are pathogenic. For instance, *PKD1_Ala2511Pro* is classified as likely benign in PKDB and VUS in VarSome. We found a carrier of *PKD1_Ala2511Pro* had diagnosis of ADPKD as did 4 family members (pedigree in **Figure 3D**), 2 of whom had prior clinical genetic testing that identified the same *PKD1* variant.

We also found ADPKD patients with *PKD1* and *PKD2* variants not reported in the Mayo PKDB. Several individuals were observed to carry the same variant (e.g., *PKD1_Gly2034Val, PKD1_Asp3304Asn, PKD1_Ser3591Phe, PKD1_Cys51Tyr* and *PKD2_Arg320Leu*). Pedigrees and abdominal imaging for representative examples are shown in **Supplemental Figure 4**. Together, these data suggest that some of these variants should be classified as likely pathogenic.

### Other cystic genes in those diagnosed with ADPKD

Finally, of the 46 individuals with ADPKD who did not have a rare variant in *PKD1* or *PKD2* (**Figure 4A**) we identified 9 patients with rare variants in one of the atypical cystic genes, including 2 with *IFT140* PTVs, 2 with *IFT140* missense variants, 4 with missense VUS in *ALG9, HNF1B*, or *PKHD1*, and one with a *GANAB* missense variant (**Figure 4B**). *GANAB*_*Asp647Val* segregates with a cystic phenotype in two families (**Figure 4C**), including 4 with mild or atypical ADPKD, 1 with multiple hypodensities in kidneys and liver, 1 with bilateral kidney cysts before age 20, and 1 with no imaging available for review. CT images for 4 carriers of *IFT40* missense variants are shown in **Supplemental Figure 5**.

**Figure 4.**
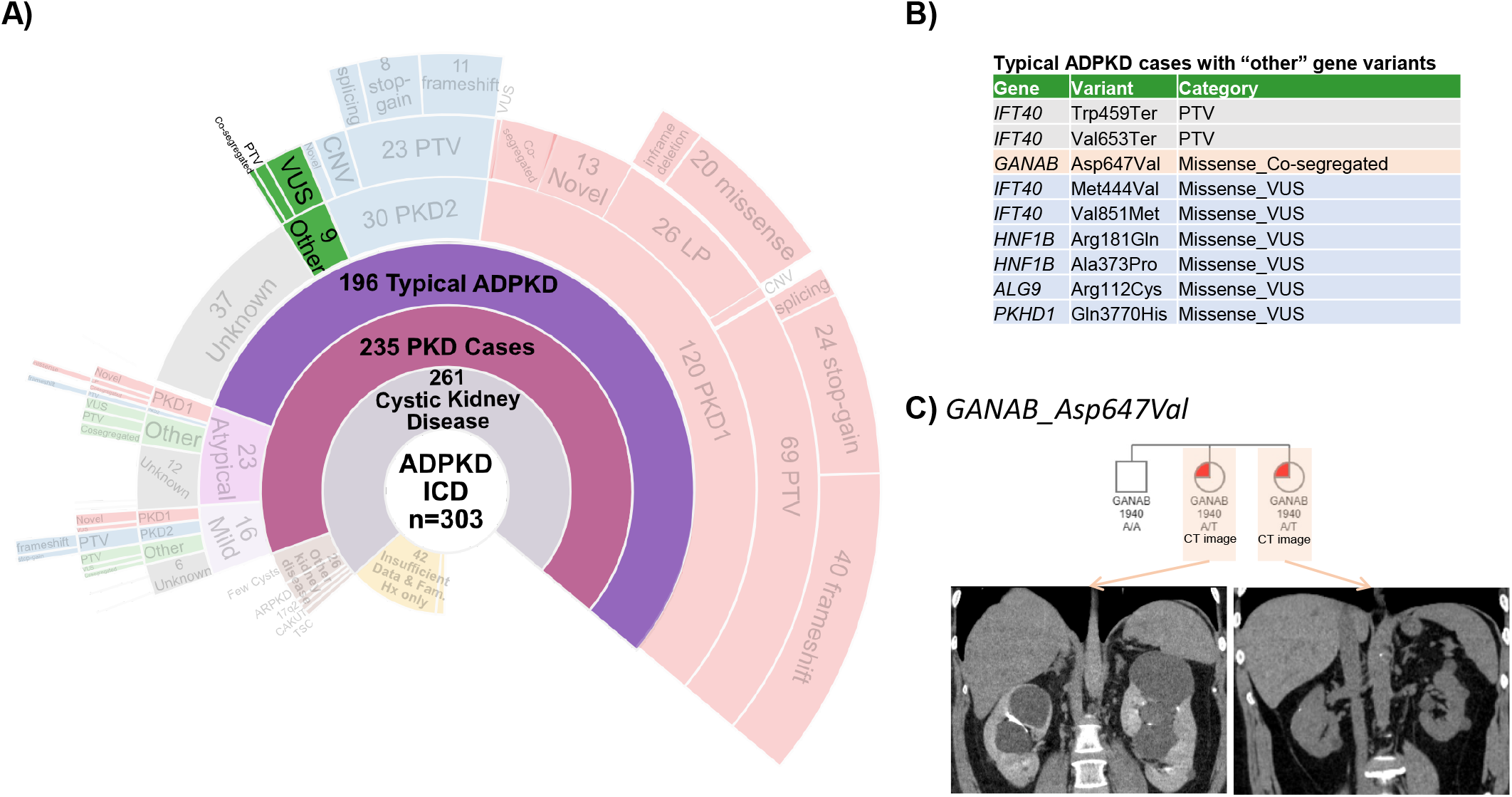
ADPKD cases with variants in genes other than PKD1/PKD2. **A)** Sunburst plot highlighting the subset of 9 ADPKD patients with no PKD1 or PKD2 variant. WES was evaluated for any variant in ALG8, ALG9, DNAJB11, GANAB, HNF1b, IFT40 LRP5, PKHD1, PRKCSH, SEC63, SEC61b. **B)** The patients had PTVs in IFT40, 6 more have a missense variant in one of these genes and one patient had a previously unreported GANAB_Asp647Val variant, **C)** Pedigrees of 2 carriers of GANAB_Asp647Val. CT Imaging from participants with GANAB_Asp647Val

## Discussion

Overall, these data show that a genotype-first approach using high quality exome sequence data can detect susceptibility to ADPKD with high positive predictive value. In the phenotype-based approach, we successfully identified rare variants in cystic genes in 180/235 (76.6%) of patients with ADPKD and 159/196 (81.1%) typical ADPKD patients, based on examination of exome sequence data and careful clinical phenotyping. We also show that CNV deletion or PTVs in atypical cystic genes are much less penetrant and associated with milder cystic phenotypes than *PKD1* or *PKD2*.

Reassuringly, we found that 94% of individuals with exome sequencing-detected PTVs or CNVs in *PKD1* had ADPKD. Thus, the vast majority of individuals who are found to have *PKD1* or *PKD2* PTV or CNV variants in high quality exome sequencing will receive a correct prediction, which could have important implications for genomic screening programs. Since penetrance for ADPKD is thought to be 100%, we estimate that ∼6% of *PKD1* PTV carriers may have a false positive finding, due to the presence of 6 large pseudogenes in *PKD1*. Therefore, confirmatory Sanger sequencing should be done to confirm a pathogenic *PKD1* variant found incidentally in asymptomatic individuals without family history.

Somewhat surprisingly, few individuals with *PKD1* or *PKD2* missense variants, previously described as likely pathogenic in the Mayo PKDB, had ADPKD. This discordance may reflect a lower penetrance of these types of variants and was also noted in a study examining the Exome Aggregation Consortium (ExAC) data, where there was 6.9-fold higher than expected rate of previously reported disease-causing variants in *PKD1* and *PKD2* (28).. Alternatively, older studies identifying these missense variants as the causal variant for ADPKD may be incorrect. For example, *PKD1_Ala2511Pro* which is classified as likely benign in the Mayo PKDB, but co-segregates with disease in one family in our cohort. Additional work using population-based cohorts, such as ours, is needed to validate pathogenicity of missense variants in *PKD1* and *PKD2*.

We also demonstrate the value of including multiple atypical cystic genes in genetic testing as several patients with ADPKD had variants in atypical cystic genes (e.g., *GANAB, IFT140, HNF1B, ALG9, PKHD1*). In genotype-based analyses several (*ALG8, ALG9, GANAB, HNF1B, IFT140, SEC63*), but not all, putative atypical cystic genes are associated with increased risk of ICD code-diagnosed cystic kidney/liver disease. However, the overall frequency of ADPKD was low in carriers of variants in these genes, and zero for many atypical ADPKD/PLD genes. Neither *LRP5* nor *PRKCSH* were associated with increased risk of kidney/liver cysts ICD codes. The full extent to which variants in atypical cystic genes (e.g., *GANAB, DNAJB11, ALG8, PRKCSH, SEC63)* cause cystic kidney or liver disease requires additional study and deeper phenotyping (i.e., imaging review) as we have done for one of the atypical cystic genes (*ALG9)* (8). Future studies are also needed to better understand the significance of mild cystic disease and whether cystic disease caused by atypical genes are responsive to therapies to slow ADPKD progression such as vasopressin receptor 2 antagonism (15).

Our data clearly show that exome sequencing is a great starting point for population-based identification of the vast majority of ADPKD patients, many of whom may be undiagnosed or unaware of their risk of developing ADPKD. A prior study found limited utility in exome sequencing *PKD1*, showing only 50% sensitivity due to issues with low read depth and sequencing quality in exons 1-33, which are also present in *PKD1P1-PKD1P6* pseudogenes (14). In our cohort, we had excellent read depth and sequencing quality throughout most of the *PKD1* gene. Out of 58 *PKD1* PTVs that we identified, 51/58 variants were correctly assigned to *PKD1;* 2 variants were misassigned to a pseudogene, 1 predicted splice site variant did not agree with spliceAI (https://spliceailookup.broadinstitute.org/), and 4 variants had either poor sequencing quality or were in a difficult to sequence G-rich region. Based on our experience effective exome capture and high-quality deep sequencing, augmented with manual sequence curation as needed for some variants, ensure the utility of exome sequencing to identify causal *PKD1* variants.

Most individuals with ADPKD have not undergone clinical genetic testing. However, new clinical genetic testing panels are becoming more common place in nephrology practice, including many that report exonic variants in tens or hundreds of genes. Our data demonstrate that most of these approaches, as well as sequencing of the entire exome in a clinic setting like ours, will have excellent positive predictive value to identify causal variants in *PKD1 and PKD2*, especially if accompanied by careful bioinformatic evaluation and Sanger sequence validation in some cases. In addition to an increase in exome sequencing services for medical genetic testing and research purposes, there has been an increase in direct-to-consumer genetic testing, including use of exome sequencing and return of results for certain actionable conditions (30). Careful curation of any of these types of exome sequencing data should facilitate early identification of at-risk individual for ADPKD.

## Data Availability

The data supporting the findings of this study are available within the article and its Supplementary Data files. Additional information for reproducing the results described in the article is available upon reasonable request and subject to a data use agreement.

## Acknowledgments

This work was supported by NIDDK106515 to A.C and GM111913 to T.M. The content is solely the responsibility of the authors and does not necessarily represent the official views of the National Institutes of Health.

## Author Contributions

Conceptualization: A.C, B.S.M., G.S., T.M.; Data curation: A.C., B.S.M, J.Z.L., B.F., S.J., Y.A., M.T., W.J.T., G.S. R.G.C.; Formal Analysis: A.C., B.S.M., J.Z.L; Funding acquisition: A.C., T.M.; Writing – original draft: A.C., B.S.M.; Writing – review & editing: A.C., B.S.M., J.Z.L., T.M.

## Ethics Declaration

This research was approved by the Geisinger Clinic Institutional Review Board and included 173,954 participants in the MyCode Health Initiative who have exome sequencing data obtained as part of the Geisinger-Regeneron DiscovEHR collaboration. All participants provided written informed consent, and all experiments were performed in accordance with relevant guidelines and regulations. The authors did not have access to any identifying information for the participants. The human phenotype and genotype data in this study were all deidentified by a “data broker” who was not involved in the study before any analysis was performed. De-identified clinical data was obtained from electronic health records (EHR).

