## Supplemental Data and Methods for "Diagnostic Utility of Exome Sequencing for Polycystic Kidney Disease"

**ARPKD, 17q12 syndrome, tuberous sclerosis, and CAKUT participants with ICD codes for PKD**

Among the 6 patients with clinical phenotype consistent with ARPKD, there were 2 related patients who had a large deletion in *PKHD1* along with *PKHD1*_Gln3407Ter, as expected with the recessive inheritance of ARPKD. One patient was heterozygous for a *PKHD1* stop gain (Arg496Ter), 1 was heterozygous for a *PKHD1* missense variant classified as pathogenic in ClinVar, and 2 had no protein-changing variants in the genes examined. Two patients had 17q12 syndrome, a large chromosomal deletion encompassing *HNF1B*, and exhibited the clinical phenotype renal cysts and diabetes syndrome (**Supplemental Table 4**). The 2 participants with a tuberous sclerosis clinical phenotype had *TSC2* variants detected. Among the 3 participants with CAKUT, 1 had a large deletion in *PKD2*, 1 had 2 rare missense variants in *FRAS1* and a rare missense variant in *SALL1*, and 1 had a pathogenic variant in *PKHD1*.

We identified 4 carriers of *PKD1_Gly2034Val*; 3 of whom had ADPKD (2 with ICD diagnosis, 1 on additional imaging review), and 1 had insufficient clinical information as she was less than 25 years of age.

Of 2 unrelated individuals with the *PKD1_Cys51Tyr* variant, 1 had ADPKD and the other individual had congenital solitary kidney with multiple cysts. The two carriers of *PKD2_Arg320Leu* are unrelated and had no known relatives, so pedigrees analysis was not possible. Both *PKD1_Cys51Tyr* and *PKD2_Arg320Leu* are classified as VUS with minor pathogenic evidence per ACMG criteria in VarSome

**Supplemental Methods**

Statistical analysis of “ADPKD” and “any kidney of liver cysts" associations

Firth’s **l**ogistic regression was used to assess the association of PTV and CNVs with ADPKD and kidney/liver cyst determined by ICD code (**Table 1**). The regression was adjusted for age, sex, year of first outpatient encounter at clinic, and genetically-determined ancestry. For these analyses, first- or second-degree relatives in the cohort were removed. For each gene, the logistic regression coefficient represents the expected change in log-odds of the outcome occurring if a PTV is present in the gene. A larger coefficient may be interpreted as a larger increase in the relative risk of the outcome (see statistical methods for regression details).

Steps to verify diagnostic accuracy

*Bioinformatic approach to validate assignment of variants to PKD1 and its pseudo genes*

Six pseudogenes on chromosome 16, *PKD1P1-PKD1P6*, replicate with high homology exons 1-33 in *PKD1*. Presence of these pseudogenes can confound interpretation of sequencing data as the sequence captured and amplified from the pseudogenes may be misaligned to *PKD1* and alternatively sequence captured from *PKD1* maybe incorrectly misaligned to one of the pseudogenes. We therefore took additional bioinformatic steps to address this potential issue. The main purpose of this exercise is to identify potential false negative variants that may have been misaligned to one of the pseudogenes in the population exome analysis. We manually forced aligned all reads from *PKD1* and pseudogenes *PKD1P1-PKD1P6* to the *PKD1* sequence and assigned variants to these forced aligned sequences which yielded 39 subject samples with 20 potential variants. While exons 1 to 33 in *PKD1* and its pseudogenes are 98.4% identical (31), there are some unique loci in almost every exon. We assumed that those unique loci were reads that belonged to *PKD1* and not indicative of a variant in a pseudogene read. Therefore, if a read included both a unique *PKD1* site and a rare variant, we classified that read as a real *PKD1* variant. Alternatively, if a read included a unique *PKD1* site but not a variant or a unique pseudogene locus with a variant, we attributed that read as a pseudogene. This process allowed us to verify that 34 participants did not have variants in *PKD1* mis-attributed to a pseudogene. The additional 5 participants’ DNA was used for long range PCR followed by Sanger sequencing. All Sanger sequences samples matched the *PKD1* reference sequence This indicates that the reads assigned to *PKD1* are truly *PKD1* reads and not from the pseudogenes.

*Use of available clinical genetic testing*

We also examined data from a convenience sample of 17 MyCode participants with cystic kidney disease or ESKD who had undergone clinical genetic testing and had either a pathogenic or VUS in a cystic gene. There was 100% concordance in this convenience sample with variants in cystic genes for all 17 participants detected using MyCode exome sequencing data (10 *PKD1* variants, 1 *HNF1B* variant, 1 *ALG8* variants, 2 *PKHD1* variants, 1 *TSC2*, 2 *PKHD1* partial deletions, 2 large *PKD1* deletions, 3 *PKD2* variants).
