## Supplemental Figures and Tables for "Diagnostic Utility of Exome Sequencing for Polycystic Kidney Disease"

### Slide 1
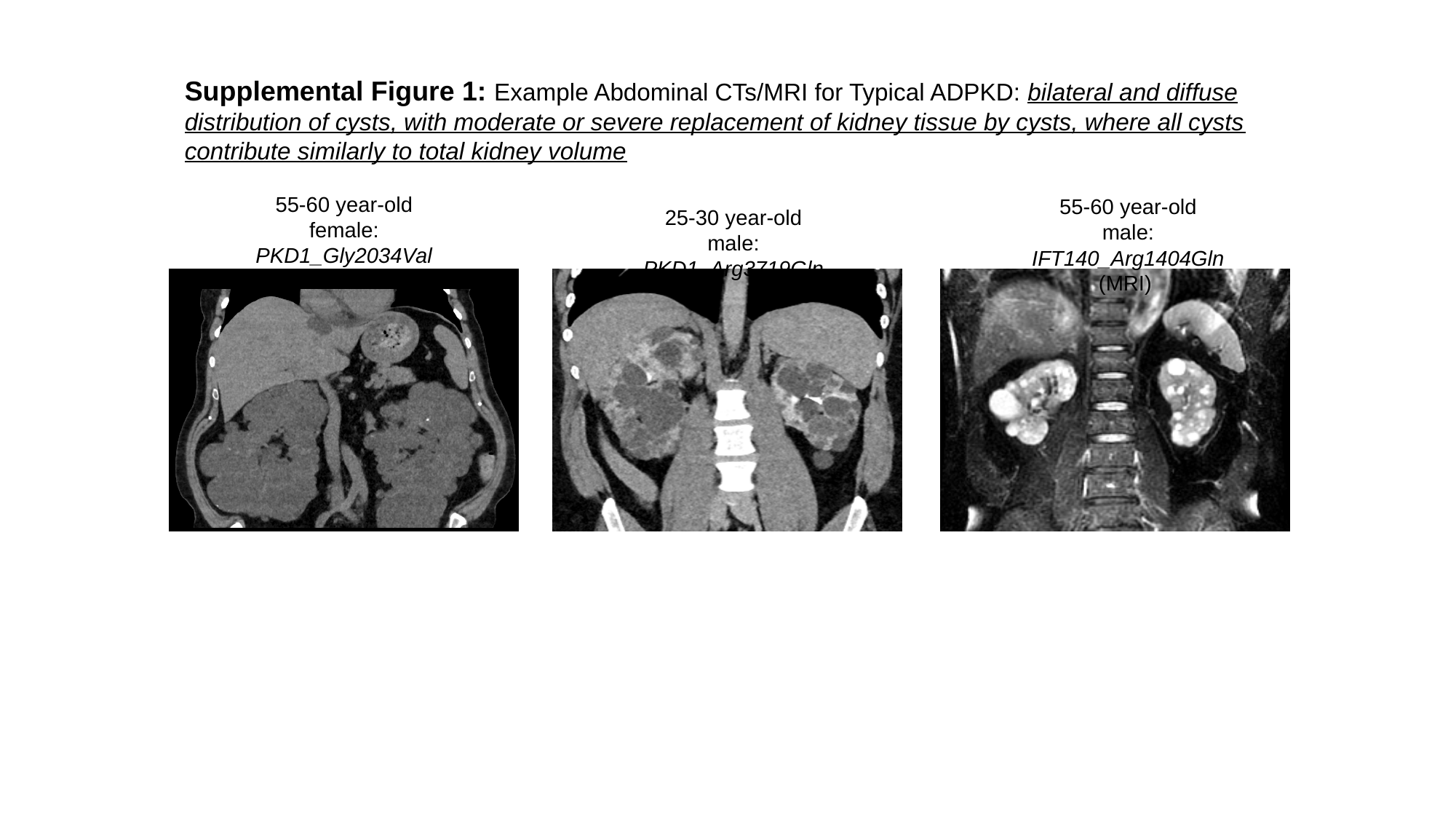

Supplemental Figure 1: Example Abdominal CTs/MRI for Typical ADPKD: bilateral and diffuse distribution of cysts, with moderate or severe replacement of kidney tissue by cysts, where all cysts contribute similarly to total kidney volume
55-60 year-old female:
PKD1_Gly2034Val
55-60 year-old male:
IFT140_Arg1404Gln
(MRI)
25-30 year-old male:
PKD1_Arg3719Gln

### Slide 2
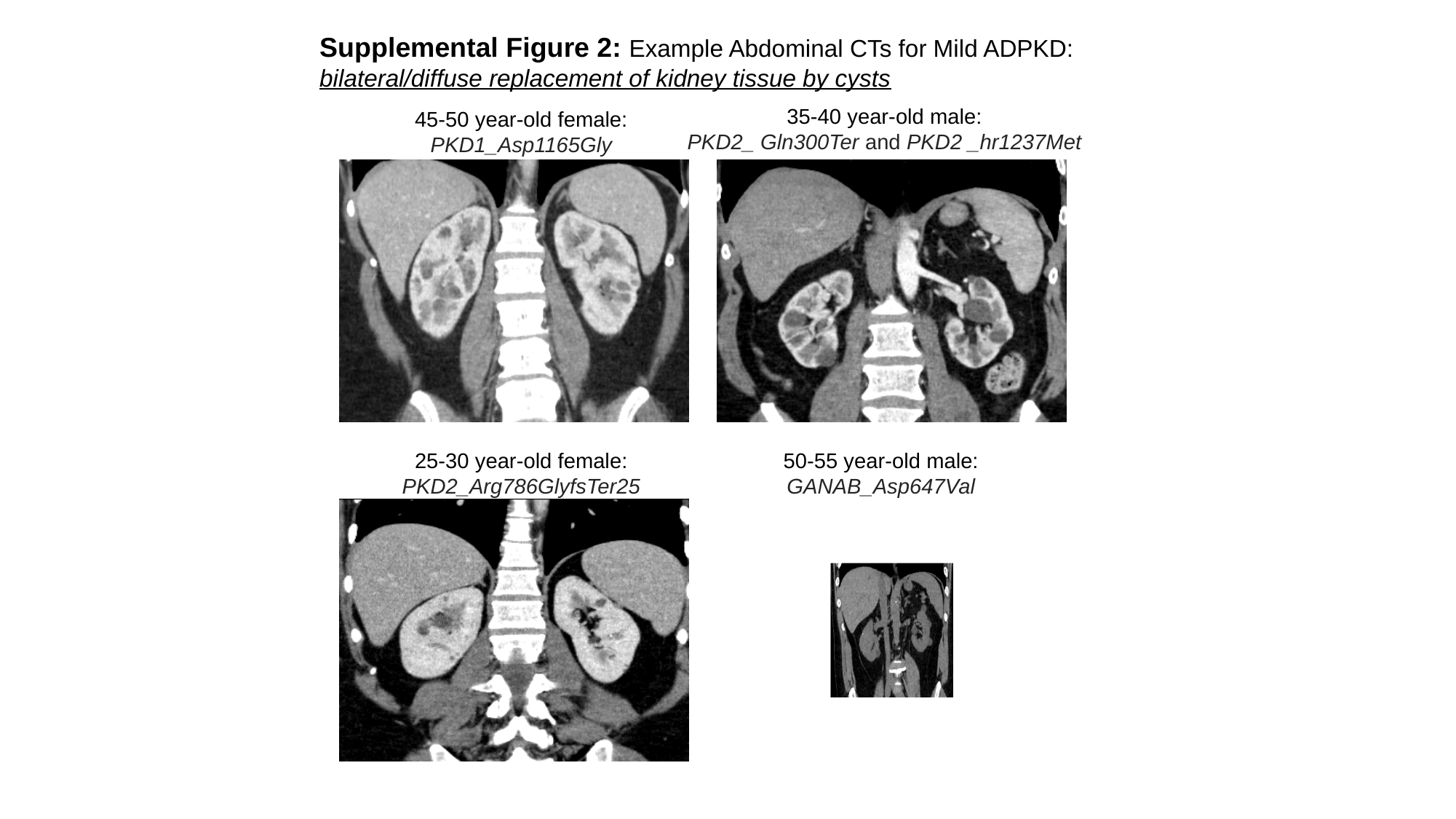

Supplemental Figure 2: Example Abdominal CTs for Mild ADPKD: bilateral/diffuse replacement of kidney tissue by cysts
35-40 year-old male:
PKD2_ Gln300Ter and PKD2 _hr1237Met
45-50 year-old female:
PKD1_Asp1165Gly
25-30 year-old female:
PKD2_Arg786GlyfsTer25
50-55 year-old male:
GANAB_Asp647Val

### Slide 3
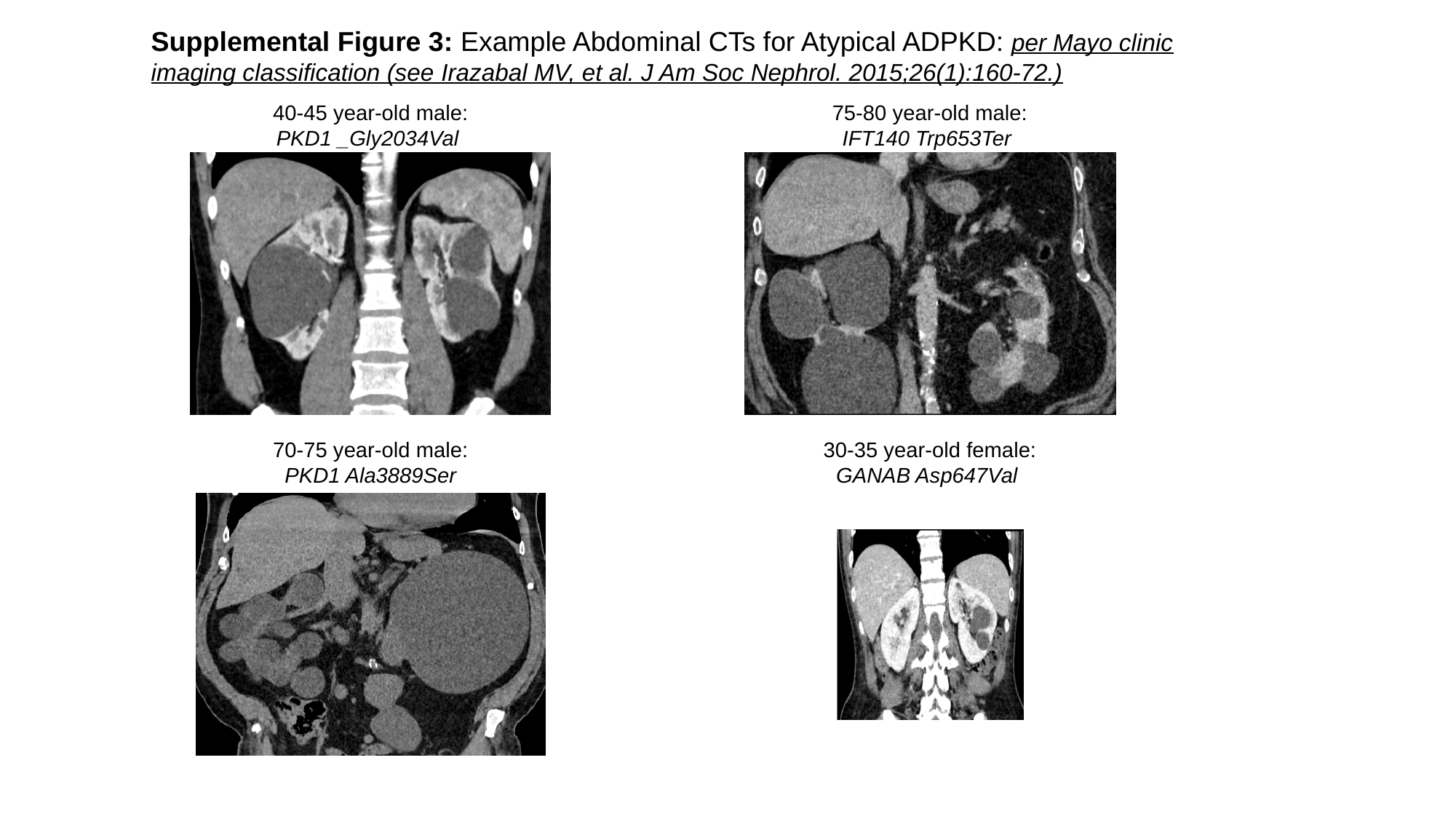

Supplemental Figure 3: Example Abdominal CTs for Atypical ADPKD: per Mayo clinic imaging classification (see Irazabal MV, et al. J Am Soc Nephrol. 2015;26(1):160-72.)
40-45 year-old male:
PKD1 _Gly2034Val
75-80 year-old male:
IFT140 Trp653Ter
70-75 year-old male:
PKD1 Ala3889Ser
30-35 year-old female:
GANAB Asp647Val

### Slide 4
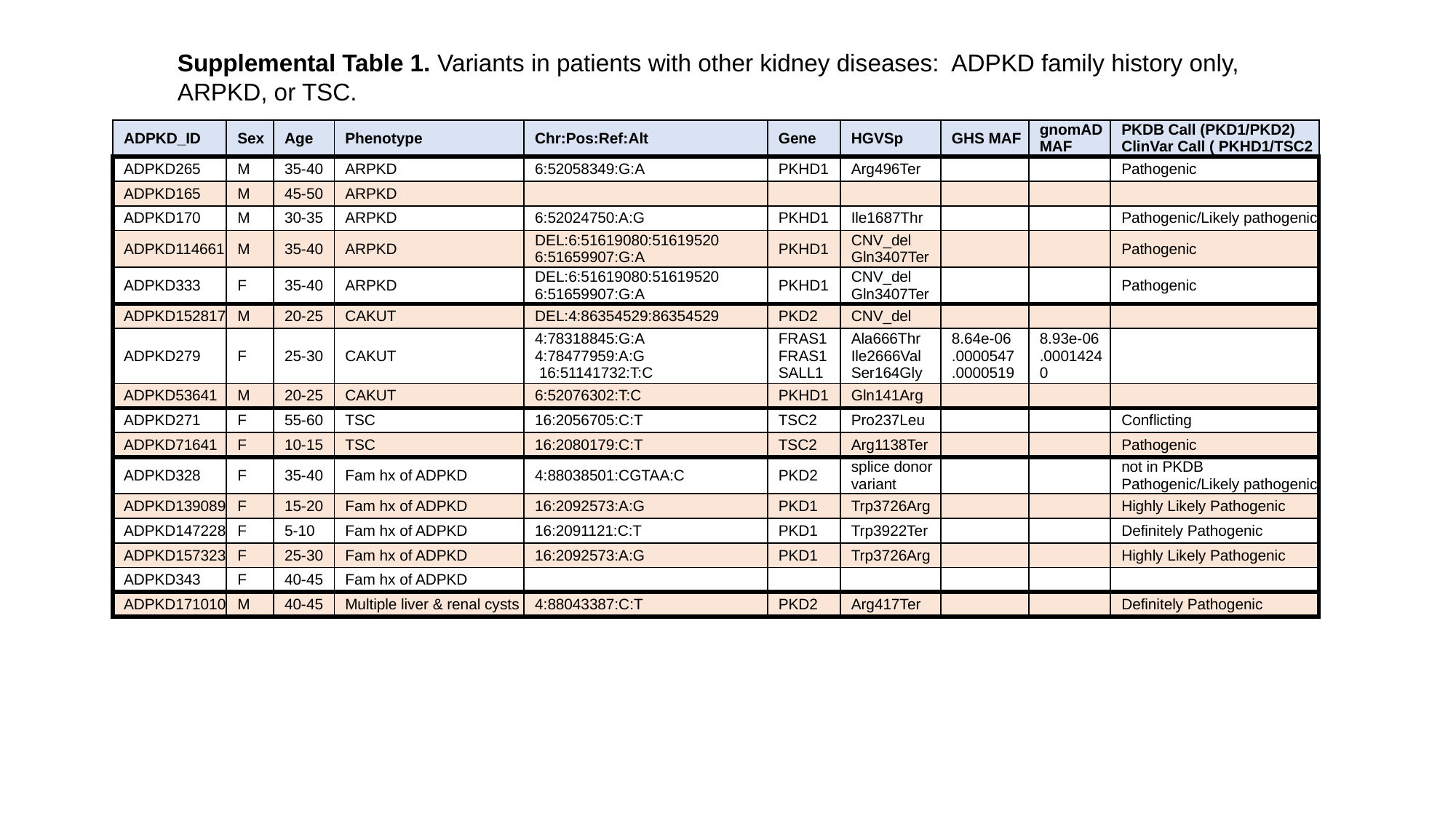

Supplemental Table 1. Variants in patients with other kidney diseases: ADPKD family history only, ARPKD, or TSC.
| ADPKD\_ID | Sex | Age | Phenotype | Chr:Pos:Ref:Alt | Gene | HGVSp | GHS MAF | gnomAD MAF | PKDB Call (PKD1/PKD2) ClinVar Call ( PKHD1/TSC2 |
| --- | --- | --- | --- | --- | --- | --- | --- | --- | --- |
| ADPKD265 | M | 35-40 | ARPKD | 6:52058349:G:A | PKHD1 | Arg496Ter | | | Pathogenic |
| ADPKD165 | M | 45-50 | ARPKD | | | | | | |
| ADPKD170 | M | 30-35 | ARPKD | 6:52024750:A:G | PKHD1 | Ile1687Thr | | | Pathogenic/Likely pathogenic |
| ADPKD114661 | M | 35-40 | ARPKD | DEL:6:51619080:51619520 6:51659907:G:A | PKHD1 | CNV\_del Gln3407Ter | | | Pathogenic |
| ADPKD333 | F | 35-40 | ARPKD | DEL:6:51619080:51619520 6:51659907:G:A | PKHD1 | CNV\_del Gln3407Ter | | | Pathogenic |
| ADPKD152817 | M | 20-25 | CAKUT | DEL:4:86354529:86354529 | PKD2 | CNV\_del | | | |
| ADPKD279 | F | 25-30 | CAKUT | 4:78318845:G:A 4:78477959:A:G 16:51141732:T:C | FRAS1 FRAS1 SALL1 | Ala666Thr Ile2666Val Ser164Gly | 8.64e-06 .0000547 .0000519 | 8.93e-06 .0001424 0 | |
| ADPKD53641 | M | 20-25 | CAKUT | 6:52076302:T:C | PKHD1 | Gln141Arg | | | |
| ADPKD271 | F | 55-60 | TSC | 16:2056705:C:T | TSC2 | Pro237Leu | | | Conflicting |
| ADPKD71641 | F | 10-15 | TSC | 16:2080179:C:T | TSC2 | Arg1138Ter | | | Pathogenic |
| ADPKD328 | F | 35-40 | Fam hx of ADPKD | 4:88038501:CGTAA:C | PKD2 | splice donor variant | | | not in PKDB Pathogenic/Likely pathogenic |
| ADPKD139089 | F | 15-20 | Fam hx of ADPKD | 16:2092573:A:G | PKD1 | Trp3726Arg | | | Highly Likely Pathogenic |
| ADPKD147228 | F | 5-10 | Fam hx of ADPKD | 16:2091121:C:T | PKD1 | Trp3922Ter | | | Definitely Pathogenic |
| ADPKD157323 | F | 25-30 | Fam hx of ADPKD | 16:2092573:A:G | PKD1 | Trp3726Arg | | | Highly Likely Pathogenic |
| ADPKD343 | F | 40-45 | Fam hx of ADPKD | | | | | | |
| ADPKD171010 | M | 40-45 | Multiple liver & renal cysts | 4:88043387:C:T | PKD2 | Arg417Ter | | | Definitely Pathogenic |

### Slide 5
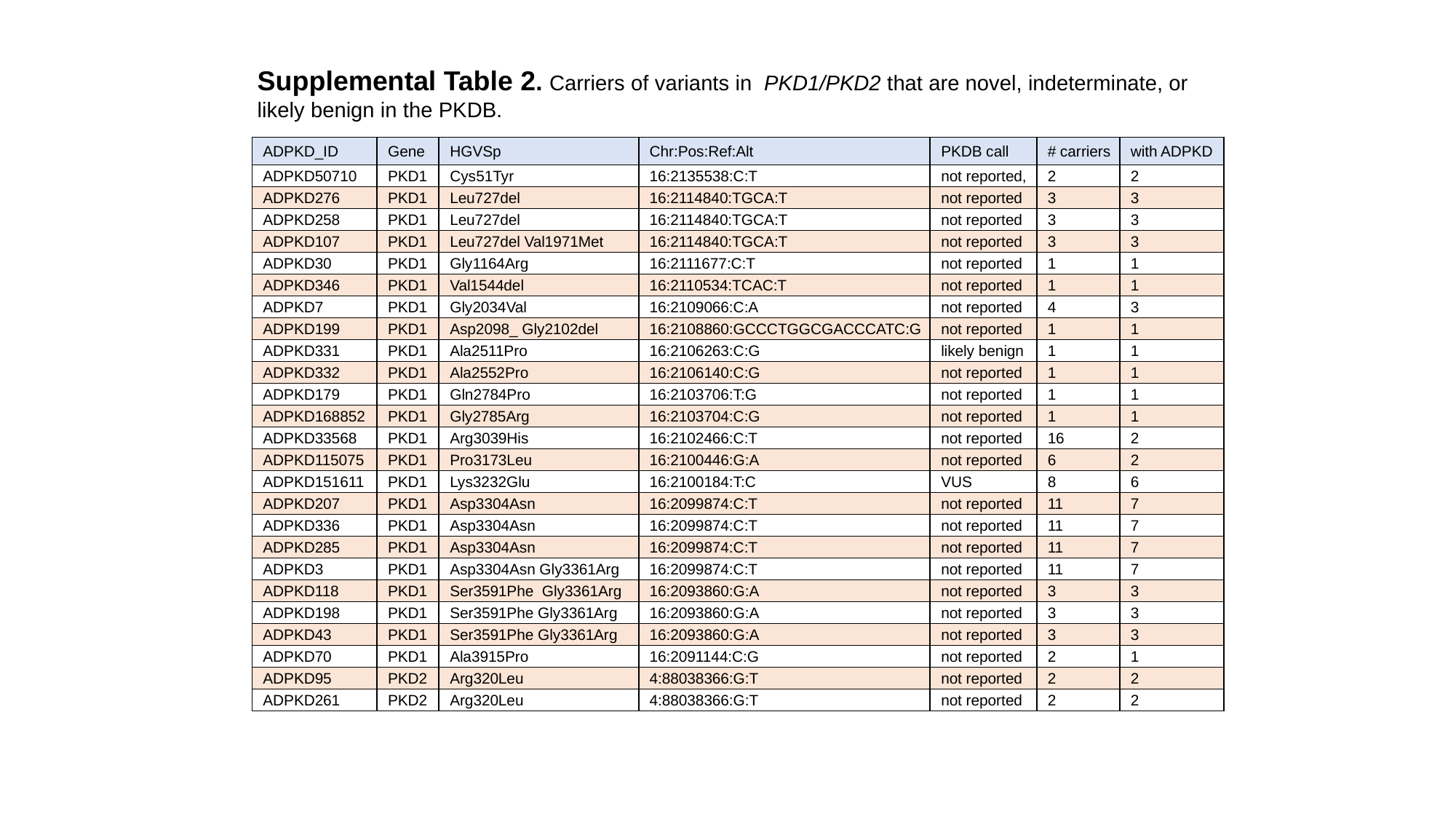

Supplemental Table 2. Carriers of variants in PKD1/PKD2 that are novel, indeterminate, or likely benign in the PKDB.
| ADPKD\_ID | Gene | HGVSp | Chr:Pos:Ref:Alt | PKDB call | # carriers | with ADPKD |
| --- | --- | --- | --- | --- | --- | --- |
| ADPKD50710 | PKD1 | Cys51Tyr | 16:2135538:C:T | not reported, | 2 | 2 |
| ADPKD276 | PKD1 | Leu727del | 16:2114840:TGCA:T | not reported | 3 | 3 |
| ADPKD258 | PKD1 | Leu727del | 16:2114840:TGCA:T | not reported | 3 | 3 |
| ADPKD107 | PKD1 | Leu727del Val1971Met | 16:2114840:TGCA:T | not reported | 3 | 3 |
| ADPKD30 | PKD1 | Gly1164Arg | 16:2111677:C:T | not reported | 1 | 1 |
| ADPKD346 | PKD1 | Val1544del | 16:2110534:TCAC:T | not reported | 1 | 1 |
| ADPKD7 | PKD1 | Gly2034Val | 16:2109066:C:A | not reported | 4 | 3 |
| ADPKD199 | PKD1 | Asp2098\_ Gly2102del | 16:2108860:GCCCTGGCGACCCATC:G | not reported | 1 | 1 |
| ADPKD331 | PKD1 | Ala2511Pro | 16:2106263:C:G | likely benign | 1 | 1 |
| ADPKD332 | PKD1 | Ala2552Pro | 16:2106140:C:G | not reported | 1 | 1 |
| ADPKD179 | PKD1 | Gln2784Pro | 16:2103706:T:G | not reported | 1 | 1 |
| ADPKD168852 | PKD1 | Gly2785Arg | 16:2103704:C:G | not reported | 1 | 1 |
| ADPKD33568 | PKD1 | Arg3039His | 16:2102466:C:T | not reported | 16 | 2 |
| ADPKD115075 | PKD1 | Pro3173Leu | 16:2100446:G:A | not reported | 6 | 2 |
| ADPKD151611 | PKD1 | Lys3232Glu | 16:2100184:T:C | VUS | 8 | 6 |
| ADPKD207 | PKD1 | Asp3304Asn | 16:2099874:C:T | not reported | 11 | 7 |
| ADPKD336 | PKD1 | Asp3304Asn | 16:2099874:C:T | not reported | 11 | 7 |
| ADPKD285 | PKD1 | Asp3304Asn | 16:2099874:C:T | not reported | 11 | 7 |
| ADPKD3 | PKD1 | Asp3304Asn Gly3361Arg | 16:2099874:C:T | not reported | 11 | 7 |
| ADPKD118 | PKD1 | Ser3591Phe Gly3361Arg | 16:2093860:G:A | not reported | 3 | 3 |
| ADPKD198 | PKD1 | Ser3591Phe Gly3361Arg | 16:2093860:G:A | not reported | 3 | 3 |
| ADPKD43 | PKD1 | Ser3591Phe Gly3361Arg | 16:2093860:G:A | not reported | 3 | 3 |
| ADPKD70 | PKD1 | Ala3915Pro | 16:2091144:C:G | not reported | 2 | 1 |
| ADPKD95 | PKD2 | Arg320Leu | 4:88038366:G:T | not reported | 2 | 2 |
| ADPKD261 | PKD2 | Arg320Leu | 4:88038366:G:T | not reported | 2 | 2 |

### Slide 6
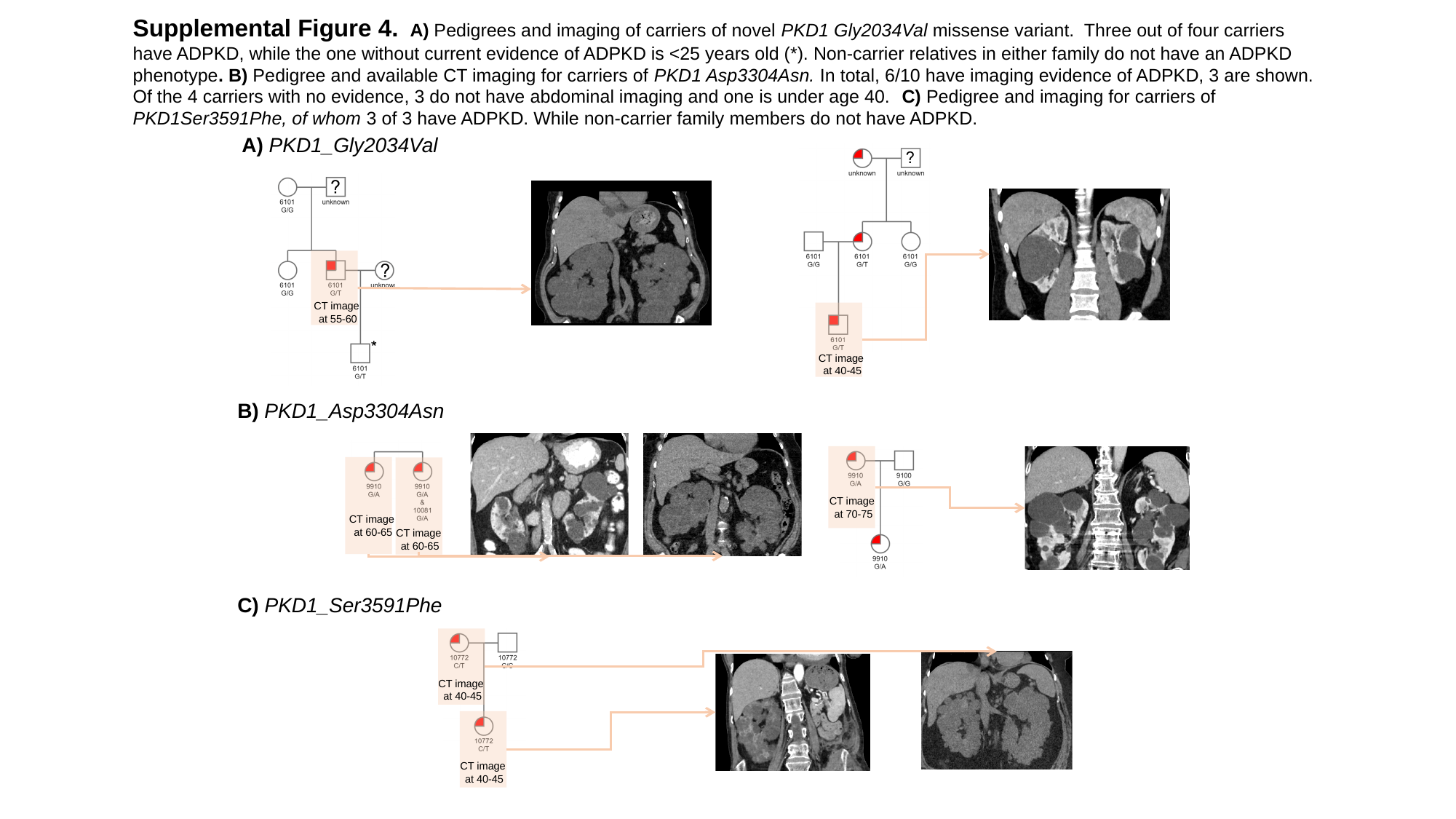

Supplemental Figure 4. A) Pedigrees and imaging of carriers of novel PKD1 Gly2034Val missense variant. Three out of four carriers have ADPKD, while the one without current evidence of ADPKD is <25 years old (*). Non-carrier relatives in either family do not have an ADPKD phenotype. B) Pedigree and available CT imaging for carriers of PKD1 Asp3304Asn. In total, 6/10 have imaging evidence of ADPKD, 3 are shown. Of the 4 carriers with no evidence, 3 do not have abdominal imaging and one is under age 40. C) Pedigree and imaging for carriers of PKD1Ser3591Phe, of whom 3 of 3 have ADPKD. While non-carrier family members do not have ADPKD.
A) PKD1_Gly2034Val
CT image
 at 55-60
CT image
 at 40-45
B) PKD1_Asp3304Asn
CT image
 at 70-75
CT image
 at 60-65
CT image
 at 60-65
C) PKD1_Ser3591Phe
CT image
 at 40-45
CT image
 at 40-45

### Slide 7
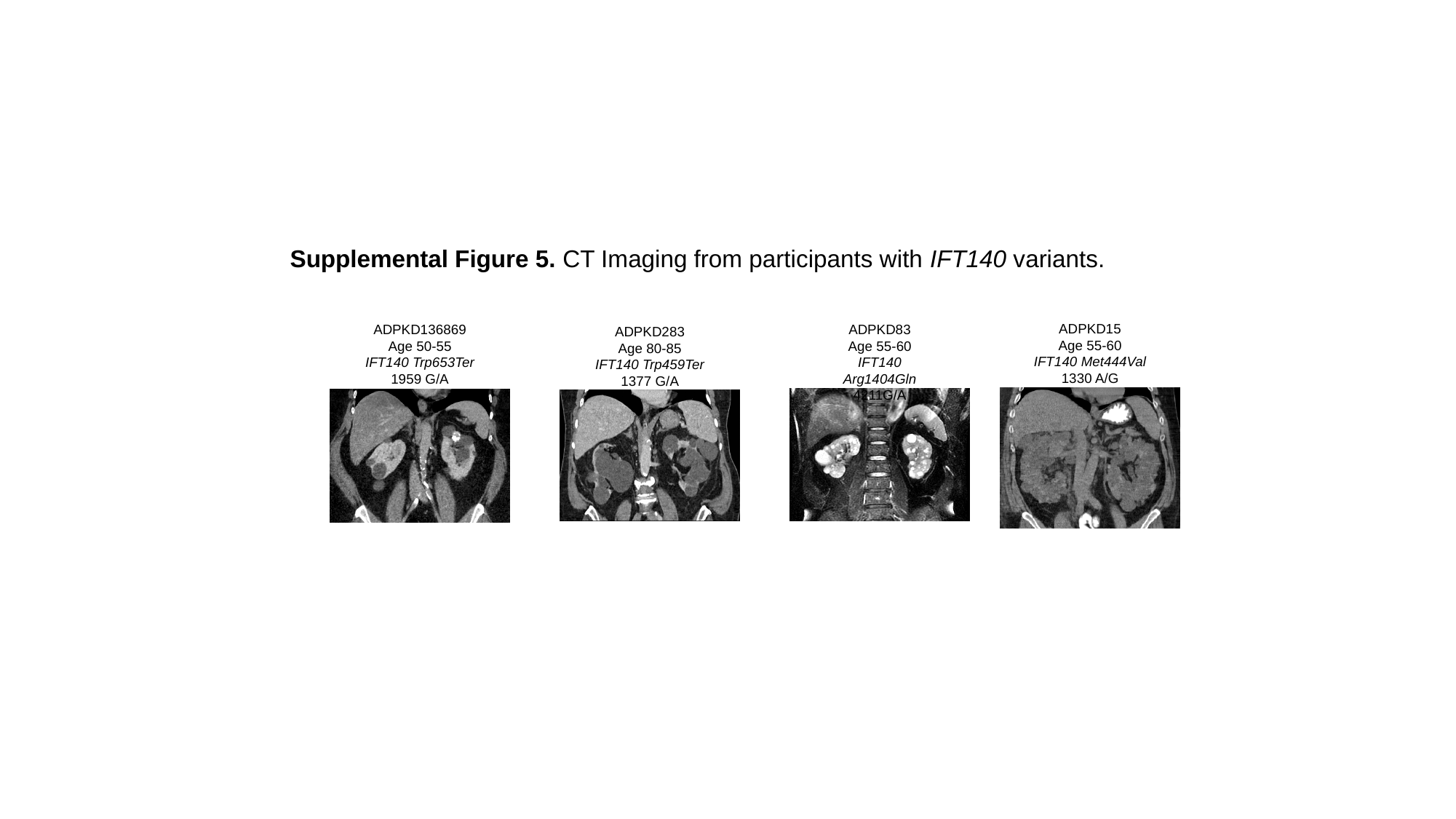

Supplemental Figure 5. CT Imaging from participants with IFT140 variants.
ADPKD15
Age 55-60
IFT140 Met444Val
1330 A/G
ADPKD83
Age 55-60
IFT140 Arg1404Gln
4211G/A
ADPKD136869
Age 50-55
IFT140 Trp653Ter
1959 G/A
ADPKD283
Age 80-85
IFT140 Trp459Ter
1377 G/A
